# Longitudinal interferon signatures are associated with disease flare in dermatomyositis

**DOI:** 10.64898/2026.09.15.26362919

**Authors:** Peixi Liu, Hsiang Chou, Martin Jarry, Marie Hudson, Sonia Del Rincon, Philippe Lefrançois, Valérie Leclair

**Affiliations:** Lady Davis Institute for Medical Research, Montreal, Canada; Interdisciplinary Cancer Sciences Program, McGill University, Montreal, QC, Canada; Department of Immunology, University of Pittsburgh School of Medicine, Pittsburgh, PA, USA; Patient representative; Division of Clinical and Translational Research, McGill University, Montreal, QC, Canada; Division of Rheumatology, Jewish General Hospital, Montreal, Canada; Division of Dermatology, Jewish General Hospital, Montreal, Canada

**Author notes:** Corresponding author: Valérie Leclair MD PhD, Division of Rheumatology, Jewish General Hospital, A.725-3755 chemin de la Côte-Sainte-Catherine, Montreal, Quebec, Canada H3T 1E2. Key Indexing Terms: Dermatomyositis, interferon, disease flare, monitoring,autoimmune diseases. Source of support: This research is funded by Arthritis Society Canada (grant #23-0000000215). Conflicts of interest: VL received honoraria from Novartis. MH received unrestricted research grants, educational grants and honoraria from Boehringer Ingelheim, Pfizer, Astra Zeneca and Merck. Statement of ethics and consent: Ethics approval for this study was obtained at the Jewish General Hospital by the Research Ethic Board of the CIUSSS du Centre-Ouest-de-l’Île-de-Montréal in Montréal (#2023-3492). All subjects provided informed written consent to participate in the study.

## Abstract

**Background:** Disease flares in dermatomyositis (DM) are unpredictable and often detected weeks to months after their onset. Interferon (IFN)-related gene signatures are implicated in DM, but their value for longitudinal flare prediction remains unclear. We hypothesized that longitudinal transcriptomic profiling using minimally invasive blood microsampling would detect IFN gene signatures preceding and during DM disease flares.

**Methods:** In this feasibility study, patients were included if they had inactive or minimally active disease and at least one of the following criteria: recent diagnosis within 2 years, a documented flare in the prior year, or ongoing tapering of immunosuppressive therapy. Patients self-assessed their disease activity weekly for 6 months using the International Myositis Assessment and Clinical Study group (IMACS) patient global assessment (PtGA; numerical rating scale, range 0-10). Disease flares, defined as a PtGA increase >2 for 2 consecutive weeks, triggered an in-person visit for confirmation by a physician. During the follow-up period, patients also self-collected weekly blood microsamples (125uL) that were shipped by courier to the study center. From those weekly blood microsamples, RNA sequencing was performed on 54 samples collected across 9 timepoints using an Illumina platform. Bioinformatic analyses included alignment, quality assessment, and gene-level quantification to evaluate differential gene expression and RNA processing changes during DM flares.

**Results:** Six DM patients were included in this study (2 anti-MDA5, 2 anti-Mi2, 1 anti-NXP2, 1 no myositis-specific antibodies). All patients were women with a mean age (SD) at study entry of 54(8) years. Three of the six patients experienced a flare during their 6-month follow-up. The mean (SD) RNA integrity score for the weekly blood microsamples collected was 7,61 (0,64). IFN scores were significantly higher in flare-associated samples than in no flare samples (linear mixed-effects model with individual patient as a random intercept, P = 5.13 × 10⁻⁴). Gene set enrichment analysis of hallmark gene sets identified enrichment of IFN-related gene sets in pre-flare and flare samples with IFN Alpha Response and IFN Gamma Response showing the highest positive normalized enrichment scores.

**Conclusion:** This study demonstrates the technical feasibility of combining home-based longitudinal blood microsampling with transcriptomic profiling in dermatomyositis. IFN gene score were higher in flare-associated than in no-flare samples, supporting further investigation of IFN-related signatures as candidate biomarkers for longitudinal disease monitoring.

## Introduction

Autoimmune inflammatory myopathies (AIM) are acquired autoimmune diseases characterized by inflammation in different organ systems such as the skin and musculoskeletal system. Dermatomyositis is one of the most frequent AIM subsets with an overall incidence of 2-15 cases/million/year and a prevalence of 4-20 cases/100 000^1^. After diagnosis, dermatomyositis patients are often treated with glucocorticoids, intravenous immunoglobulins (IVIG) and at least one glucocorticoid-sparing agent (e.g., methotrexate, mycophenolate mofetil). Remission may be achieved in ∼60% of patients by 1 year, but disease flares are frequent after remission^2,3^. Disease flares often require glucocorticoids to be re-introduced, can lead to increased organ damage and are associated with increased healthcare costs.

Traditional monitoring for disease activity in dermatomyositis consists of clinical assessments and laboratory tests such as levels of muscle enzymes every 3-6 months. However, this approach often identifies flares several weeks or even months after their onset. Thus, in the absence of reliable monitoring strategies for early flare detection, many clinicians hesitate to de-escalate immunosuppression to avoid accrual of organ damage. To improve disease monitoring in AIM, several novel biomarkers have been studied. However, those biomarkers remain challenging to implement in clinical practice. For example, interferon (IFN) related proteins (e.g., CXCL11) and gene scores have shown associations with AIM disease activity^4–6^. Still, existing assays to measure IFN (e.g., proteins, gene expression) are not standardized nor accessible to clinicians and were never studied to assess their utility for disease flare detection^7,8^.

Longitudinal analysis of blood transcriptional profiles using serial fingerstick blood specimens have been used to predict flares in rheumatoid arthritis^9^. Serial fingerstick blood specimens have the potential to capture pre-flare biological data and is a promising avenue for personalized medicine in the field of inflammatory diseases. We aimed to assess the feasibility of remote patient monitoring in dermatomyositis using patient-reported outcomes, blood microsamples, and transcriptomics. We hypothesized that dermatomyositis remote patient monitoring would be feasible and that IFN gene expression would be enriched during and in the weeks preceding clinical flares of dermatomyositis.

## METHODS

### Study population

Participants for this feasibility study were identified from the Canadian Inflammatory Myopathy Study (CIMS) registry. Patients enrolled in CIMS are followed longitudinally with standardized assessments and self-administered questionnaires including the International Myositis Assessment and Clinical Study (IMACS) group core set measures^10^. The IMACS core set measures include the physician and patient global assessments (PtGA/PhGA, visual analog scale (VAS); range 0-10), extramuscular assessment (VAS; range 0-10), manual muscle testing (MMT8, range 0-150), Health Assessment Questionnaire (HAQ, range 0-3), and muscle enzyme levels. Ethics approval for the study was obtained at the Jewish General Hospital by the Research Ethic Board of the CIUSSS du Centre-Ouest-de-l’Île-de-Montréal in Montréal (#2023-3492). All subjects provided informed written consent to participate in the study.

### Remote patient monitoring

Included patients at risk of flares were followed longitudinally for 6 months with collection of weekly PtGA measurements and weekly blood microsamplings. At 0, 3 and 6 months (and as needed if experiencing an intercurrent flare), patients were seen in person by a study investigator for disease activity assessments. Participants performed microsampling of 3 drops of blood (i.e.,125 uL) at home using tubes prefilled with 375uL of stabilizing reagent (RNA*later*, Thermo Fischer). The frozen samples were transported weekly with ice packs by courier for ribonucleic acid (RNA) extraction at the Lady Davis Institute (Montreal, Canada). RNA was extracted using the RiboPure RNA blood purification kit (Thermo Fischer) and purified accounting for the small volume of blood^11^. RNA quality and quantity was assessed using the Agilent 2100 BioAnalyzer system that provides RNA integrity number (RIN) scores ranging from 1 to 10, with higher scores indicating less degradation and higher integrity. Library preparation was done using the Illumina TruSeq mRNA stranded Library kit and sequencing using the HiSeq2500 Illumina system at the McGill Genome Center with an average sequencing depth of 55 million read. RNA sequencing was performed on samples collected at 0, 3 and 6 months and those corresponding to a flare including the 3 weeks preceding the onset of the flare (i.e., pre-flare) provided a RIN score ≥4. At the end of the study, participants completed an acceptability questionnaire to share their perspective on the microsampling method used ^12^ .

### Disease flare definition

A disease flare was defined as an increase of ≥2 points from a previous recorded visit on the weekly IMACS PtGA for ≥2 consecutive weeks which was then confirmed by a study physician as a flare if also corresponding to an increase of ≥2 points on the IMACS PhGA^13^. This flare definition aligns with consensus definitions used in clinical trials^13,14^. The flare onset date was considered to be the date when the patient first indicated a PtGA increase of ≥2 points.

### RNA sequencing data processing

Raw paired-end RNA sequencing reads were assessed using FastQC (v0.12.1), and quality metrics across samples were summarized using MultiQC (v1.25.1)^15,16^. Adapter sequences and low-quality bases were removed using fastp, followed by post-trimming quality assessment using FastQC and MultiQC^17^. Trimmed reads were aligned to the human reference genome (GRCh38) using HISAT2 (v2.2.1)^18^. Alignment files were sorted and indexed using SAMtools, and gene-level read counts were generated using HTSeq-count (v2.0.4) with Ensembl gene annotations^19,20^. Gene-level count files were generated for downstream analyses.

### Differential gene expression analysis

Differential gene expression analysis was performed in R using DESeq2^21^. Raw gene counts were normalized using the median-of-ratios method. Samples collected during the pre-flare and flare periods were combined into a flare-associated group and compared with samples collected during no-flare periods. Individual study identifiers were included in the DESeq2 design to account for repeated longitudinal sampling. Differentially expressed genes were defined using an adjusted P value < 0.1, absolute log₂ fold change ≥ 0.5, and mean normalized expression (baseMean) > 10. Variance-stabilizing transformation (VST) was applied for visualization and exploratory analyses. Principal component analysis (PCA) was performed using VST-transformed expression values, and volcano plots were generated using ggplot2^22^.

### Gene set enrichment analysis

Gene set enrichment analysis (GSEA) was performed using the fgsea package in R^23^. Genes were ranked according to the DESeq2 Wald statistic from the comparison between flare-associated and no-flare samples. Hallmark gene sets from the Molecular Signatures Database (MSigDB) were used for pathway enrichment analysis. Normalized enrichment scores (NES), nominal P values, and Benjamini–Hochberg-adjusted P values were calculated. Because of the exploratory nature of this feasibility study, Hallmark gene sets with an adjusted P value < 0.2 were considered enriched.

### IFN score calculation

Type I IFN activity was quantified using a previously reported 10-gene interferon-stimulated gene (ISG) signature consisting of *IFI27*, *IFI44L*, *IFIT1*, *ISG15*, *RSAD2*, *SIGLEC1*, *LY6E*, *MX1*, *USP18*, and *OAS1*, using a modified approach adapted from Baechler et al.^24^ For each gene, DESeq2-normalized counts were log₂-transformed, and the median expression across all no-flare samples was used as the reference. Relative expression values for each gene were calculated with respect to this reference, and the IFN score for each sample was defined as the median relative expression across the 10 ISGs. Differences in IFN scores between no-flare and flare-associated samples were evaluated using a linear mixed-effects model, with flare status as a fixed effect and individual identifier as a random intercept to account for repeated longitudinal measurements. Models were fitted using the lme4 package, and P values were calculated using Satterthwaite’s approximation implemented in lmerTest^25,26^.

### Transcript isoform analysis

Transcript-level abundance was quantified using Salmon (v1.10.2) and imported into R using tximport. Isoform switching between flare and no flare samples was evaluated using IsoformSwitchAnalyzeR with the DEXSeq-based differential isoform usage framework. Patient identity was included in the DEXSeq design matrix to account for repeated longitudinal sampling. Lowly expressed transcripts and transcripts with low isoform fractions were filtered before differential isoform usage analysis. Differential isoform usage was evaluated using differential isoform fraction (dIF) together with false discovery rate-adjusted P values.

### Statistical analyses

Statistical analyses were performed in R. Differential gene expression was assessed using DESeq2, pathway enrichment was performed using fgsea, and differences in IFN scores were evaluated using linear mixed-effects models implemented in lme4 and lmerTest. For transcriptome-wide analyses, P values were adjusted for multiple testing using the Benjamini–Hochberg false discovery rate procedure unless otherwise specified. Analysis-specific significance thresholds are described above.

## RESULTS

### Population

Six patients with dermatomyositis at risk of flares were included in this feasibility study (Table 1). All patients were women with a mean age (SD) at study entry of 54(8) years. All but one patient had positive myositis-specific antibody (2 anti-MDA5, 2 anti-Mi2, 1 anti-NXP2). All were on at least one immunosuppressor when they entered the study, and 4 were on tapering doses of glucocorticoids. Three patients (50%) flared during their follow-up. Two of them were on tapering doses of prednisone, while a third one was on stable immunosuppression but had been tapered off intravenous immunoglobulins before study enrollment.

**Table 1.** Characteristics of patients with dermatomyositis included in the study.

|  | Patient 1 | Patient 2 | Patient 3 | Patient 4 | Patient 5 | Patient 6 |
| --- | --- | --- | --- | --- | --- | --- |
| **MSA** | Anti-NXP2 | Anti-Mi2 | Anti-MDA5 | None | Anti-Mi2 | Anti-MDA5 |
| **MAA** | Anti-Ro52 | - | Anti-Ro52, -<br>Ku | - | - | - |
| **Clinical features at diagnosis** |  |  |  |  |  |  |
| **Rash** | + | + | + | + | + | + |
| **Calcinosis** | + | - | - | - | - | - |
| **Raynaud** | + | - | + | + | - | + |
| **Arthritis** | - | + | + | - | - | - |
| **ILD** | - | - | + | - | - | + |
| **Dysphagia** | + | - | - | + | - | - |
| **Muscle weakness** | + | + | + | + | + | + |
| **Max CK (U/L)** | 2722 | 7132 | 321 | 480 | 681 | 252 |
| **Malignancy** | - | - | Breast | - | - | - |
| **Clinical features at month 0** |  |  |  |  |  |  |
| **Immunosuppression** | HCQ, TOF | HCQ, MMF | TAC, HCQ | MTX | MTX | HCQ, TAC, TOF |
| **Prednisone dose** | 0 | 10 mg | 12.5 mg | 15 mg | 20 mg | 0 |
| **Immunosuppression change during follow-up** | None | Pred taper | Pred taper | Pred taper | Pred taper | None |
| **PhGA (range 0-10)** | 3 | 5 | 2 | 3 | 3 | 4 |
| **PtGA (range 0-10)** | 0 | 2 | 2 | 1 | 1 | 1 |
| **Extramuscular VAS (range 0-10)** | 0 | 2 | 1 | 1 | 0 | 2 |
| **MMT8, 0-150** | 150 | 141 | 150 | 150 | 142 | 150 |
| **CK level (U/L)\*** | 180 | 98 | 31 | 16 | 124 | 129 |
| **Ferritin (ug/L)\*** | 101 | 12 | 62 | 98 | N/A | 49 |
| **Clinical features at flare** |  |  |  |  |  |  |
| **Manifestations** | No flare | No flare | No flare | Rash, alopecia, myositis, dysphagia | Arthralgia, myalgias, rash | Rash |
| **Immunosuppression** | - | - | - | MTX | MTX | HCQ, TAC, TOF |
| **Prednisone dose** | - | - | - | 0 | 8 mg | 0 |

*Abbreviations:* MSA, myositis-specific antibodies; MAA, myositis-associated antibodies; ILD, interstitial lung disease; CK, creatine kinase; HCQ, hydroxychloroquine; TOF, tofacitinib; MMF, mycophenolate mofetil; TAC, tacrolimus; MTX, methotrexate; PhGA, physician global assessment; PtGA, patient global assessment; MMT8, manual muscle testing. \*Within 3 months of baseline visit

### Remote patient monitoring

A total of 132 weekly patient questionnaires and blood microsamples were delivered by courier to the study team. Longitudinal PtGA and disease flares temporality are shown in Figure 2. Four out of 6 patients completed their acceptablility questionnaires and reported that the procedure was acceptable to them, did not interfere with their usual activities and that they felt confident performing it (Supplementary Table 1). From the blood microsamples received (n=132), 7 had undetectable RIN (5%), and 3 had RIN <4 (2%).

**Figure 1.**
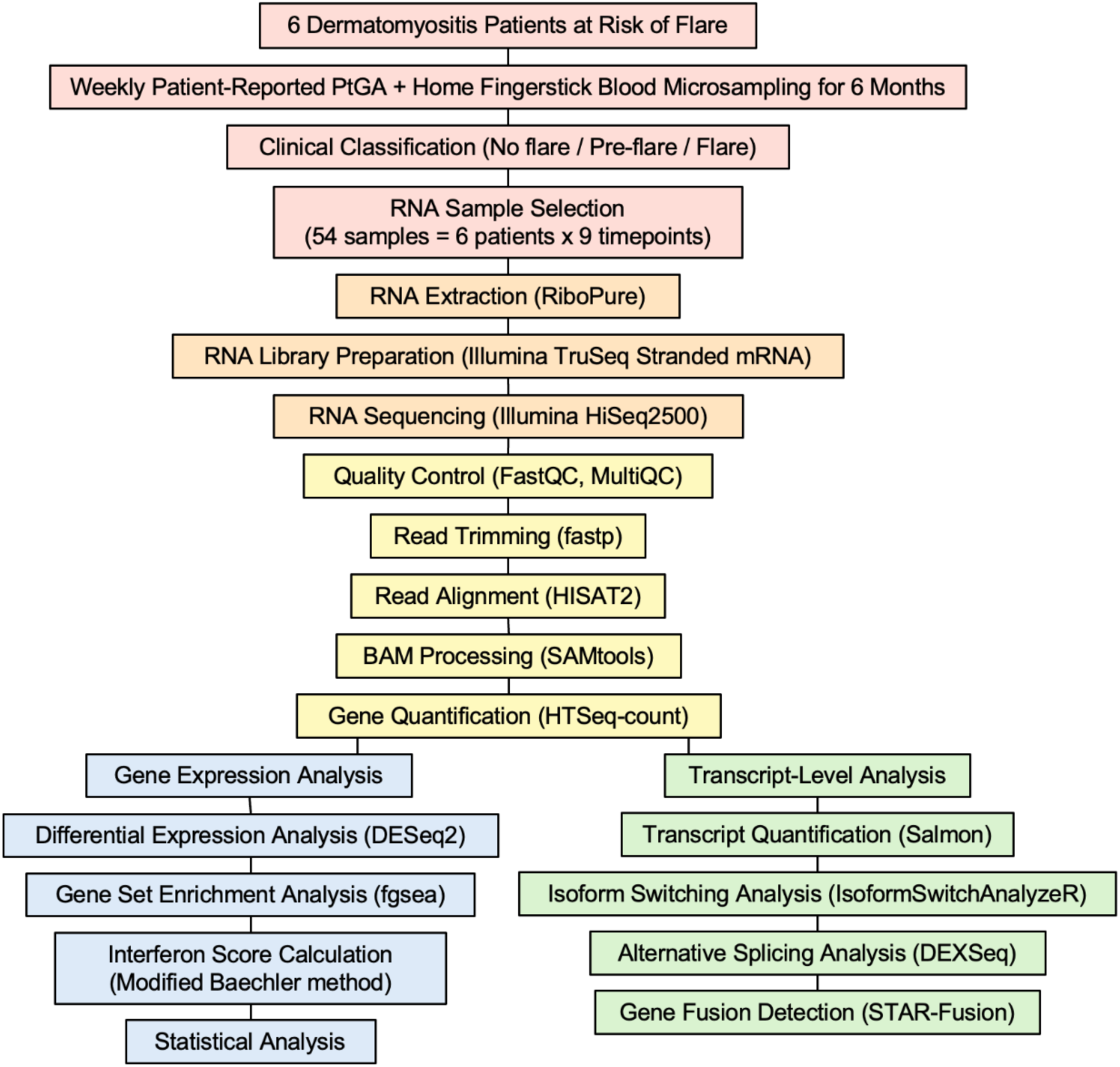
Overview of the study design and workflow, from patient monitoring and blood microsampling to RNA sequencing and downstream bioinformatic analyses.

**Figure 2.**
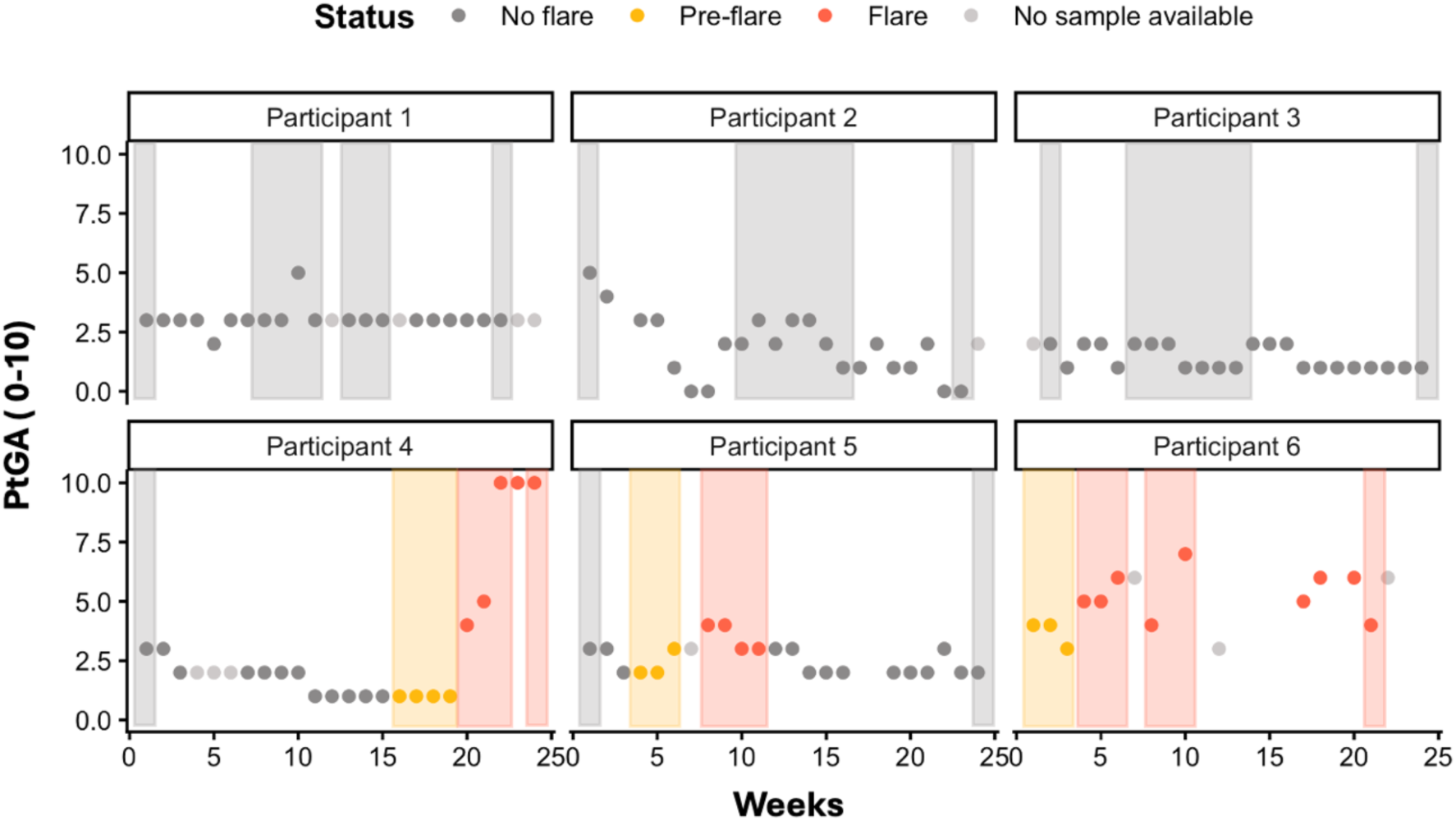
RNA sample selection and longitudinal disease activity. Longitudinal disease activity measured weekly using the IMACS Patient Global Assessment (PtGA; NRS 0-10) during the 6-month follow-up period. The pre-flare (3-4 weeks preceding a flare) and the flare periods are indicated by yellow and red, respectively. For participants 1, 2 and 3 who did not flare, RNA samples were randomly selected from their follow-up period. In total, 54 RNA samples were included in the study, comprising 10 pre-flare, 14 flare and 30 no-flare samples.

For the microsamples with RIN >4 (n=122, 92%), the mean RIN (SD) was 7,61 (0,64) (Supplementary Figure 1). A total of 54 blood samples were selected for RNA sequencing and transcriptomic analyses (Figure 2). For all participants, baseline and last visit were analyzed. For those who flared, pre-flare samples corresponding to the 3-4 weeks before the identification of the flare, depending on samples availability, were selected in addition to samples collected during the flare. For patients that did not flare, RNA samples were randomly selected to assess for possible variation in transcriptomic profiles despite disease quiescence.

### Longitudinal IFN scores

Longitudinal IFN scores were calculated using a previously published IFN-stimulated gene signature^24^. IFN scores showed substantial inter-patient heterogeneity but were generally higher during pre-flare and flare periods than during no-flare periods (Figure 3A). When pre-flare and flare samples were combined into a “flare” group, IFN scores were significantly higher in flare-associated samples than in no flare samples (linear mixed-effects model with individual patient as a random intercept, P = 5.13 × 10⁻⁴; Figure 3B). Individual longitudinal trajectories showed temporal variation in IFN activity around flare events. Because pre-flare and flare samples were combined in the primary analysis, predictive performance was not evaluated.

**Figure 3.**
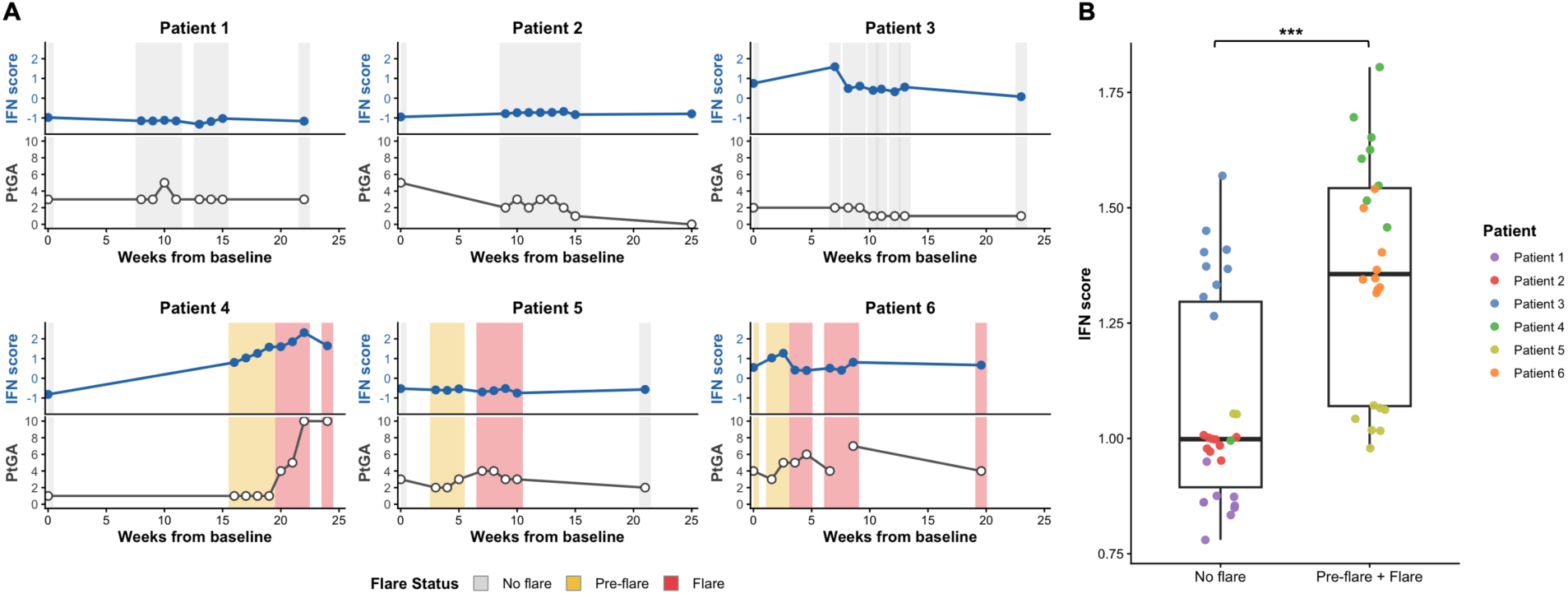
Longitudinal changes in IFN gene score and PtGA during remote monitoring of patients with dermatomyositis. **(A)** Longitudinal IFN scores (top) and patient global assessment (PtGA; bottom) for each of the six patients over the 6-month monitoring period. Grey, yellow, and red shaded regions indicate samples classified as no flare, pre-flare, and flare, respectively. Patients 1-3 did not experience a disease flare during follow-up, whereas patients 4-6 developed at least one flare. IFN scores are shown as median fold-change relative to the no-flare reference. **(B)** Comparison of IFN gne scores between no flare samples and pre-flare/flare samples. Each point represents one RNA sequencing sample and is colored by patient. Boxes indicate the interquartile range (IQR), center lines represent the median, and whiskers extend to 1.5 × IQR. Groups were compared using a linear mixed-effects model with patient as a random effect. ***P = 5.13 × 10⁻⁴.

### Transcriptomic profiles

Principal component analysis (PCA) of variance-stabilized transcriptomic profiles demonstrated substantial inter-individual variation across the longitudinal samples. Samples clustered primarily by patient, highlighting patient-specific transcriptional heterogeneity (Figure 4A). Differential expression analysis comparing pre-flare/flare samples with no flare samples identified a small number of differentially expressed genes (adjusted P < 0.1, |log₂FC| ≥ 0.5). (Figure 4B). These results indicate that although relatively few genes reached statistical significance in this longitudinal cohort, disease flare was associated with detectable transcriptomic alterations. Gene set enrichment analysis of Hallmark gene sets identified enrichment of interferon-related gene sets in pre-flare and flare samples. IFN Alpha Response and IFN Gamma Response showed the highest positive normalized enrichment scores (Figure 4C). In contrast, pathways related to heme metabolism, UV response, and reactive oxygen species showed negative enrichment. Together, these findings suggest that the pre-flare and flare periods are associated with activation of interferon-driven transcriptional programs.

**Figure 4.**
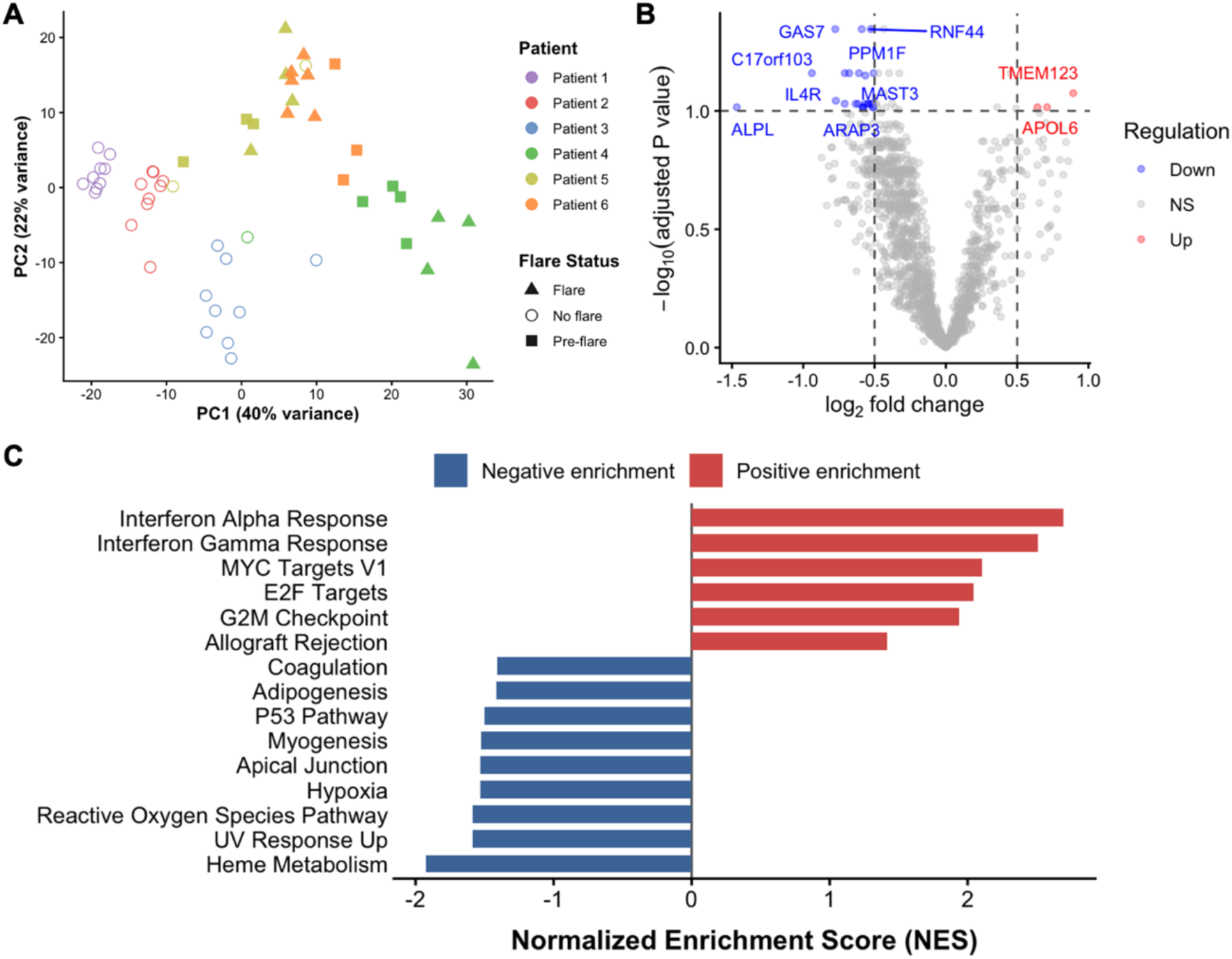
Transcriptomic changes associated with disease flare in dermatomyositis. **(A)** PCA of all RNA-seq samples colored by patient and shaped by flare status. **(B)** Volcano plot showing differentially expressed genes between flare/pre-flare and no-flare samples. Dashed lines indicate the thresholds for differential expression (|log₂ fold change| ≥ 0.5 and adjusted P < 0.1). **(C)** Hallmark GSEA based on the ranked differential expression results. Positive normalized enrichment scores (NES) indicate pathways enriched in pre-flare/flare samples, whereas negative NES indicates pathways enriched in no flare samples.

## DISCUSSION

Most studies looking at novel disease monitoring strategies in inflammatory myopathies focused on the use of wearables^27–29^, or patient reported outcomes administered digitally. In this study, we demonstrate for the first time the feasibility of using patient-reported outcomes and home-based blood microsampling and showed that the interventions were well accepted by dermatomyositis patients and generated RNA of sufficient quality for transcriptomic profiling. Moreover, IFN gene scores generated using the transcriptomic data were found to be higher in samples collected during flare periods compared to no flare periods. These findings support an association between longitudinal IFN activity and clinically defined disease flare and provide preliminary evidence that IFN-related signatures could be potential candidate biomarkers for early flare detection in dermatomyositis.

Type I IFN pathway activation is a well-established hallmark of dermatomyositis^30^. However, previous studies exploring IFN levels or gene expression in relation to disease activity relied on samples collected cross-sectionally or over few numbers of timepoints, resulting in a limited view of the dynamic changes in IFN type I activation especially around disease flare periods. A 2012 study by Reed et al. assessed IFN gene scores in 51 dermatomyositis patients (21 juvenile, 30 adults) at 2 separate timepoints and found that changes in IFN scores were correlated with changes in disease activity measures such as the IMACS PhGA^6^. More recently, a 2023 study by Tabata et al. looked at the correlations between clinical features including autoantibody status and disease activity parameters in adult dermatomyositis^31^. This study included 74 patients with >2 blood samples (153 visit pairs), although individual temporal trends or disease flares where not reported. Their results nonetheless revealed a correlation between skin disease activity and type 1 IFN gene scores that was stronger in anti-MDA5 positive patients compared to other dermatomyositis subsets. Our study provides a much deeper understanding of the dynamic changes occurring in pre-flare and flare periods in dermatomyositis patients, but also in period of quiescence which has never been reported.

By applying a previously reported IFN-stimulated gene signature to weekly home-collected blood samples for a 6-month follow-up period, our study revealed generally higher IFN scores during pre-flare and flare periods. While our differential gene expression analysis identified relatively few significantly dysregulated genes, the hallmark gene set enrichment analysis showed enrichment of IFN alpha and gamma response pathways in pre-flare and flare samples. These pathway-level changes aligned with the IFN score, indicating activation of IFN-driven transcriptional activity during dermatomyositis flare. The limited number of differentially expressed genes were expected due to the small cohort size and inter-patient variation observed in the PCA. However, our findings highlight the temporality and variability of the IFN signature in dermatomyositis warranting larger scale validation studies.

Interestingly, one patient who experienced a flare in our study had no change in her IFN gene score. Although it is difficult based on one patient to draw any conclusion, it raises interesting questions about the use of IFN signature to monitor disease activity in the spectrum of dermatomyositis disease, and in particular for change in disease activity of possibly lower magnitude which characterizes early disease flares. This patient was anti-Mi2 positive, and in the Tabata et al. study previously mentioned, anti-Mi2 positive patients were more likely to have low type 1 IFN (i.e., IFN score <2.5)^31^. That low type 1 IFN group represented a quarter of their cohort (n=46), and of those, 35% had moderate to severe skin activity suggesting that for some dermatomyositis patients, type 1 IFN gene score might not be an ideal disease activity biomarker. This supports the exploration of alternative transcriptomic signatures which might be more sensitive to detect early disease flares.

This feasibility study has some limitations. First, only six patients were included which might not represent the whole spectrum of DM patients while limiting statistical power and preventing subgroup analyses by autoantibody subtypes. Moreover, the longitudinal transcriptomic profiles clustered primarily by patient, highlighting substantial heterogeneity between individuals. However, the results generated are essential to inform the design of larger remote patient monitoring studies.

## CONCLUSION

This study demonstrates the technical feasibility of combining home-based longitudinal blood microsampling with transcriptomic profiling in dermatomyositis. IFN gene score were higher in flare-associated than in no-flare samples, supporting further investigation of IFN-related signatures as candidate biomarkers for longitudinal disease monitoring. Larger prospective studies are needed to establish temporal relationships with flare, validate discriminatory performance, and determine clinical utility.

## Data Availability

All data produced in the present study are available upon reasonable request to the authors.

## References

1. Khoo T, Lilleker JB, Thong BY, Leclair V, Lamb JA, Chinoy H. Epidemiology of the idiopathic inflammatory myopathies. Nat Rev Rheumatol. Nov 2023;19(11):695–712. doi:10.1038/s41584-023-01033-0

2. Espinosa-Ortega F, Holmqvist M, Dastmalchi M, Lundberg IE, Alexanderson H. Factors Associated With Treatment Response in Patients With Idiopathic Inflammatory Myopathies: A Registry-Based Study. Arthritis Care Res (Hoboken). Mar 2022;74(3):468–477. doi:10.1002/acr.24498

3. Tsuji H, Espinosa-Ortega F, Garcia IP, Dastmalchi M, Lundberg IE, Lodin K. Relapse Rate After Glucocorticoid-free Remission in patients with Idiopathic Inflammatory Myopathies and Validation of the International Myositis Assessment and Clinical Studies Group (IMACS) Criteria for Complete Clinical Response and Worsening. Arthritis Rheumatol. Nov 10 2025;doi:10.1002/art.43429

4. Walsh RJ, Kong SW, Yao Y, et al. Type I interferon-inducible gene expression in blood is present and reflects disease activity in dermatomyositis and polymyositis. Arthritis Rheum. Nov 2007;56(11):3784–92. doi:10.1002/art.22928

5. Greenberg SA, Higgs BW, Morehouse C, et al. Relationship between disease activity and type 1 interferon- and other cytokine-inducible gene expression in blood in dermatomyositis and polymyositis. Genes Immun. Apr 2012;13(3):207–13. doi:10.1038/gene.2011.61

6. Reed AM, Peterson E, Bilgic H, et al. Changes in novel biomarkers of disease activity in juvenile and adult dermatomyositis are sensitive biomarkers of disease course. Arthritis Rheum. Dec 2012;64(12):4078–86. doi:10.1002/art.34659

7. Rodriguez-Carrio J, Burska A, Conaghan PG, et al. Association between type I interferon pathway activation and clinical outcomes in rheumatic and musculoskeletal diseases: a systematic literature review informing EULAR points to consider. RMD Open. Mar 2023;9(1)doi:10.1136/rmdopen-2022-002864

8. Rodriguez-Carrio J, Burska A, Conaghan PG, et al. 2022 EULAR points to consider for the measurement, reporting and application of IFN-I pathway activation assays in clinical research and practice. Ann Rheum Dis. Jun 2023;82(6):754–762. doi:10.1136/ard-2022-223628

9. Orange DE, Yao V, Sawicka K, et al. RNA Identification of PRIME Cells Predicting Rheumatoid Arthritis Flares. N Engl J Med. Jul 16 2020;383(3):218–228. doi:10.1056/NEJMoa2004114

10. Miller FW, Rider LG, Chung YL, et al. Proposed preliminary core set measures for disease outcome assessment in adult and juvenile idiopathic inflammatory myopathies. Rheumatology (Oxford). Nov 2001;40(11):1262–73. doi:10.1093/rheumatology/40.11.1262

11. Robison EH, Mondala TS, Williams AR, Head SR, Salomon DR, Kurian SM. Whole genome transcript profiling from fingerstick blood samples: a comparison and feasibility study. BMC Genomics. Dec 17 2009;10:617. doi:10.1186/1471-2164-10-617

12. Sekhon M, Cartwright M, Francis JJ. Development of a theory-informed questionnaire to assess the acceptability of healthcare interventions. BMC Health Serv Res. Mar 1 2022;22(1):279. doi:10.1186/s12913-022-07577-3

13. Rider LG, Aggarwal R, Machado PM, et al. Update on outcome assessment in myositis. Nat Rev Rheumatol. May 2018;14(5):303–318. doi:10.1038/nrrheum.2018.33

14. Aggarwal R, Charles-Schoeman C, Schessl J, et al. Trial of Intravenous Immune Globulin in Dermatomyositis. N Engl J Med. Oct 6 2022;387(14):1264–1278. doi:10.1056/NEJMoa2117912

15. Ewels P, Magnusson M, Lundin S, Kaller M. MultiQC: summarize analysis results for multiple tools and samples in a single report. Bioinformatics. Oct 1 2016;32(19):3047–8. doi:10.1093/bioinformatics/btw354

16. Andrews S. FastQC: A Quality Control Tool for High Throughput Sequence Data [Online]. 2010. http://www.bioinformatics.babraham.ac.uk/projects/fastqc/

17. Chen S, Zhou Y, Chen Y, Gu J. fastp: an ultra-fast all-in-one FASTQ preprocessor. Bioinformatics. Sep 1 2018;34(17):i884–i890. doi:10.1093/bioinformatics/bty560

18. Kim D, Paggi JM, Park C, Bennett C, Salzberg SL. Graph-based genome alignment and genotyping with HISAT2 and HISAT-genotype. Nat Biotechnol. Aug 2019;37(8):907–915. doi:10.1038/s41587-019-0201-4

19. Li H, Handsaker B, Wysoker A, et al. The Sequence Alignment/Map format and SAMtools. Bioinformatics. Aug 15 2009;25(16):2078–9. doi:10.1093/bioinformatics/btp352

20. Putri GH, Anders S, Pyl PT, Pimanda JE, Zanini F. Analysing high-throughput sequencing data in Python with HTSeq 2.0. Bioinformatics. May 13 2022;38(10):2943–2945. doi:10.1093/bioinformatics/btac166

21. Love MI, Huber W, Anders S. Moderated estimation of fold change and dispersion for RNA-seq data with DESeq2. Genome Biol. 2014;15(12):550. doi:10.1186/s13059-014-0550-8

22. Wickham H. ggplot2: Elegant Graphics for Data Analysis. Springer-Verlag New York; 2016. https://ggplot2.tidyverse.org.

23. Subramanian A, Tamayo P, Mootha VK, et al. Gene set enrichment analysis: a knowledge-based approach for interpreting genome-wide expression profiles. Proc Natl Acad Sci U S A. Oct 25 2005;102(43):15545–50. doi:10.1073/pnas.0506580102

24. Baechler EC, Bauer JW, Slattery CA, et al. An interferon signature in the peripheral blood of dermatomyositis patients is associated with disease activity. Mol Med. Jan-Feb 2007;13(1-2):59–68. doi:10.2119/2006-00085.Baechler

25. Bates D, Mächler M, Bolker B, Walker S. Fitting Linear Mixed-Effects Models Using lme4. Journal of Statistical Software. 2015;67(1):1–48. doi:10.18637/jss.v067.i01.

26. Kuznetsova A, Brockhoff P, Christensen R. lmerTest Package: Tests in Linear Mixed Effects Models. Journal of Statistical Software. 82(13):1–26. 10.18637/jss.v082.i13

27. Rockette-Wagner B, Saygin D, Moghadam-Kia S, et al. Reliability, validity and responsiveness of physical activity monitors in patients with inflammatory myopathy. Rheumatology (Oxford). Dec 1 2021;60(12):5713–5723. doi:10.1093/rheumatology/keab236

28. Landon-Cardinal O, Bachasson D, Guillaume-Jugnot P, et al. Relationship between change in physical activity and in clinical status in patients with idiopathic inflammatory myopathy: A prospective cohort study. Semin Arthritis Rheum. Oct 2020;50(5):1140–1149. doi:10.1016/j.semarthrit.2020.06.014

29. Oldroyd AGS, Krogh NS, Dixon WG, Chinoy H. Investigating characteristics of idiopathic inflammatory myopathy flares using daily symptom data collected via a smartphone app. Rheumatology (Oxford). Nov 28 2022;61(12):4845–4854. doi:10.1093/rheumatology/keac161

30. Bolko L, Jiang W, Tawara N, et al. The role of interferons type I, II and III in myositis: A review. Brain Pathol. May 2021;31(3):e12955. doi:10.1111/bpa.12955

31. Tabata MM, Hodgkinson LM, Wu TT, et al. The Type I Interferon Signature Reflects Multiple Phenotypic and Activity Measures in Dermatomyositis. Arthritis Rheumatol. Oct 2023;75(10):1842–1849. doi:10.1002/art.42526

