## Supplemental files for "Longitudinal interferon signatures are associated with disease flare in dermatomyositis"

**Supplementary Figure 1**. RIN values for the microsamples collected

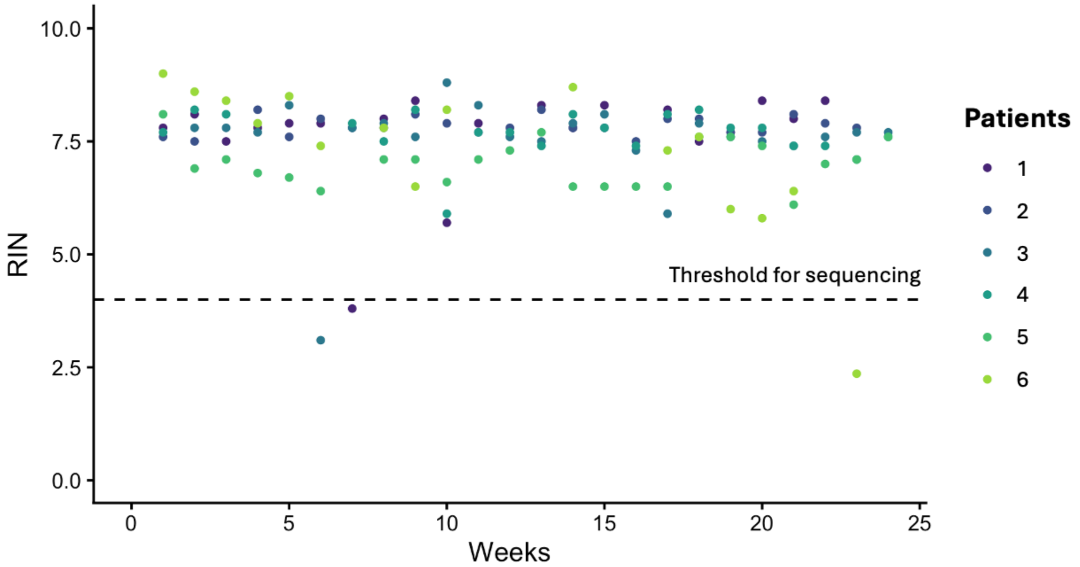


*Abbreviations*: RIN, RNA integrity number

**Supplementary Table 1**. Acceptability of blood microsampling procedure

| | Patient 1 | Patient 2 | Patient 3 | Patient 4 |
|---|---|---|---|---|
| **Microsampling** | | | | |
| How comfortable did you feel with the microsampling procedure? | Like | No opinion | Like | Strongly like |
| How much effort did the procedure take? | A little effort | No effort at all | A little effort | A little effort |
| How confident did you feel with the procedure? | Confident | Confident | Very confident | Very confident |
| The procedure Interfered with my other priorities? | Strongly disagree | No opinion | Disagree | Strongly disagree |
| How acceptable was the procedure to you? | Completely acceptable | Acceptable | Completely acceptable | Completely acceptable |

Legend: Four of the 6 participants of the feasibility study returned their acceptability questionnaire. Positive feedback in a gradient of green. Negative feedback in a gradient of orange.
